# GROMTools: scalable individual-level GReX imputation for mega-biobank-scale cohorts

**DOI:** 10.64898/2026.04.27.26351635

**Authors:** Marios Anyfantakis, Sanan Venkatesh, Jamie J. R. Bennett, Gabriel E. Hoffman, Panos Roussos, Georgios Voloudakis

## Abstract

**Motivation:** The computational burden of individual-level genetically regulated gene expression (GReX) imputation has risen sharply with the growth of human mega-biobanks and the rapid expansion of transcriptomic imputation models across tissues and single-cell hierarchies. Existing tools were not designed for this setting and require complex, memory-intensive workflows that are poorly matched to shared and cloud-based compute environments, where runtime, memory, and I/O directly determine cost and throughput. GROMTools is an open-source C++ engine with an R interface that exploits sparse prediction weights, streams PLINK2 genotypes, and writes compact binary outputs for scalable individual-level GReX imputation.

**Results:** In UK Biobank (UKBB), benchmarks on chromosome 1 across 50,000-450,000 individuals, 388,017 variants, and 11,724 gene-tissue pairs from 32 single-cell models, GROMTools produced near-identical predictions to PrediXcan and PLINK2, with minimum Pearson correlation >0.999 and maximum RMSE <0.001 across all of the imputed genes, while reducing CPU time by about 100-fold and peak memory by about 33-fold. These gains make routine biobank-scale individual-level GReX imputation practical and cost-efficient on standard CPU infrastructure.

**Availability:** GROMTools is freely available at https://github.com/voloudakislab/gromtools under the GPL v3 license. Documentation available at https://voloudakislab.github.io/gromtools/. All of our coding scripts used for quality control of data and benchmarking pipelines and all of our log files are archived at DOI: https://doi.org/10.5281/zenodo.19547333.

## Introduction

Transcriptome-wide association studies (TWAS) and related transcriptome-informed analyses have become a central framework in human complex trait genetics. These methods link germline genetic variation to gene expression and downstream phenotypes through genetically regulated gene expression (GReX). Beginning with PrediXcan (Gamazon et al., 2015) and FUSION (Gusev et al., 2016), they train expression prediction models in reference transcriptome panels and apply the resulting cis-genetic weights to summary- and individual-level biobank genotypes, typically as weighted linear combinations of local variants. This framework enables GReX imputation from genotype data and, in summary-statistics settings, direct gene-trait association testing without matched expression measurements in the target cohort. Across a wide range of complex traits and diseases, these approaches have yielded reproducible and biologically interpretable findings (Huckins et al., 2020; Voloudakis et al., 2022; Venkatesh et al., 2024).

Yet the scale of individual-level GReX imputation has increased substantially. Large human biobanks now include millions of genotyped participants, while reference prediction models continue to expand across tissues (Gamazon et al., 2015; Zhang et al., 2019) and increasingly fine single-cell hierarchies (Zeng et al., 2024; Venkatesh et al., 2024). Consequently, individual-level GReX imputation increasingly now requires aggregating sparse genetic weights across thousands of genes, many tissue and cell-type models, and very large cohorts. Existing tools were not designed for this setting and therefore depend on complex, memory-intensive workflows deployed in high-performance or cloud computing environments. In shared and fee-for-service settings, runtime, memory use, and I/O thus directly affect both cost and throughput (McDonald et al., 2012; Krissaane et al., 2020).

To address this bottleneck, we developed GROMTools (Genetically Regulated Omics tools), an open-source GReX imputation engine implemented in C++ with an R interface, designed to address this bottleneck. GROMTools exploits sparse prediction weights, streams PLINK2 genotype data efficiently, and writes compact binary outputs, enabling scalable individual-level GReX imputation without proportionate increases in memory usage. Using incremental subsampling of the UK Biobank (Sudlow et al., 2015), we benchmarked GROMTools against PrediXcan and PLINK2 and observed concordant GReX predictions together with substantial gains in CPU runtime and memory efficiency.

## Methods

GReX is a large sparse linear prediction problem. Let *Y* = *G* × *B*, where *G* ∈ ℝ ^*n*×*m*^ is the genotype matrix for *n* individuals and *m* variants, *B* is the sparse matrix of model-specific variant weights across *k* gene-model pairs, and *Y* ∈ ℝ ^*n*×*k*^ is the resulting matrix of individual-level imputed expression values. At biobank scale, the main challenge is computational rather than statistical: naive implementations typically load large genotype matrices into memory and apply general dense matrix operations that do not exploit the sparsity of *B*. GROMTools preserves the standard linear GReX model, but improves its implementation by streaming genotype data in SNP chunks and restricting computation to nonzero weights, thereby reducing memory usage, avoiding repeated scans of large genotype files, and accelerating matrix multiplication.

### Input 1 - genotype data

Before computation, the current workflow requires a prior QC step in which weight-file predictors are matched to genotype variants using chromosome-position-reference-alternate (CPRA), with complementary-allele matches permitted and effect-allele alignment resolved; full details are described elsewhere (Venkatesh et al., 2024) and we provide a companion repository (`voloudakislab / gromtools_QC_helper`) that implements this approach. Following this QC step, GROMTools maps genotype variants from the `.PVAR` manifest to the corresponding weight-file predictors for imputation. This pre-indexing removes repeated parsing and lookup from the inner computation loop. To reduce genotype access overhead, GROMTools streams PLINK2 binary files (`pgen`/`psam`/`pvar`) directly through the vendored pgenlib library. Genotypes are read in SNP chunks rather than loading the full genotype matrix into memory, and optional sample subsetting is applied before computation so that downstream buffers are sized only to the analyzed individuals. Together, these design choices reduce both memory use and input overhead, improving scalability to large cohorts.

### Input 2 - GReX prediction weights

Prediction weights are supplied as tab-separated files, one per transcriptomic imputation model, containing variant identifiers, effect alleles, and weight coefficients. At startup, GROMTools loads all weights and organizes them into a unified sparse representation indexed by variant. Because the full weight matrix spanning *M* variants and *K* gene-tissue pairs is highly sparse, this representation remains compact and allows each variant to traverse only the gene-model pairs with nonzero weights, avoiding unnecessary scans over unrelated predictors.

### Core computation

The core computation is implemented in C++ as sparse linear updates over the weight structure, accumulating GReX predictions blockwise across samples. For each variant, only the connected gene-model pairs are visited, and the corresponding prediction vectors are updated using the C-interface to the Basic Linear Algebra Subprograms (BLAS) library for efficient dense accumulation. To improve throughput, updates are further decomposed into row chunks across samples, allowing computation to proceed over contiguous slices of the genotype and output vectors in a cache-friendly manner. Although Intel Math Kernel Library (MKL) can also be used as an alternative backend where available. In the benchmarks reported here, MKL was not enabled for GROMTools, so as to provide a conservative estimate of performance under a standard non-optimized BLAS configuration.

### Output and streaming

To control memory usage, GROMTools avoids materializing the full output matrix in memory. Instead, it allocates dense output columns only when a gene-tissue pair is first touched during computation, accumulates contributions while that column remains active, and flushes completed columns to disk once all contributing variants have been processed. Export is performed in batches to a preallocated .grom file in contiguous column-major layout. As a result, peak memory depends primarily on the number of simultaneously active output columns, together with the current genotype chunk, rather than on the full cohort-by-gene matrix, thereby improving scalability to very large cohorts on standard CPU nodes.

### Software implementation

GROMTools is implemented in C++17 and exposed through an R interface using Rcpp. Genotype I/O uses vendored PLINK2/pgenlib code through `RPgenReader.cpp`, so an external pgenlib library is not required. The package is built with a standard R package toolchain using `configure` and `Makevars`. A pre-built Docker container image (`marvany/ubuntu-r422-PLINK2:1.1`) is provided for reproducible execution.

GROMTools exposes two user-facing wrapper functions, `grom_impute()` and `grom_read()`. In its standard use case, `grom_impute()` requires as input a tab-delimited weights table (`weights_path`) containing the columns `chromosome`, `ancestry`, `model_ID`, `gene`, `rsid`, and `weight`, together with `pgen_dir`, the directory containing the corresponding PLINK2 genotype files (`.pgen`, `.pvar`, `.psam`). The `chromosome` field in the weights table is expected to use the `chr*` convention (e.g. `chr1`), which is used to map each set of gene-model weights to the corresponding chromosome-specific PLINK2 file. The genotype files are therefore likewise expected to be chromosome split and contain the literal `chr*` string in their filenames. To prevent erroneous chromosome assignment, GROMTools performs internal checks to ensure exact chromosome matching and to avoid ambiguous mappings such as `chr1` being incorrectly assigned to `chr11`. Additional quality-control procedures are applied throughout the mapping process, and the resulting mappings to the `.pvar` and `.pgen` files are written to the `meta/` directory for optional user inspection.

The function further requires `grom_pfx`, which specifies the prefix of the output files. These outputs include `grom_pfx.grom`, a binary matrix containing the imputed gene-expression values for all model-gene pairs across all chromosomes; `grom_pfx.gid`, which stores the mapping of output columns to model-gene identifiers and serves a role analogous to the `.pvar` file in PLINK2; and `grom_pfx.sid`, which stores the mapping of output rows to samples and is analogous to the `.psam` file.

The companion function, `grom_read()`, provides access to previously generated GROMTools output and takes `grom_pfx` as its primary input. It also exposes optional arguments for selective loading of models, genes, or individuals, enabling targeted access to subsets of the imputed matrix without requiring full import of all results.

All remaining arguments of `grom_impute()` and `grom_read()` are optional. Detailed descriptions of these parameters, together with usage examples, are provided in the package documentation and in the `vignettes/getting-started.md` available in the GitHub repository.

### Benchmark design

We benchmarked GROMTools against PrediXcan and PLINK2 `--score` using UK Biobank genotype data. To assess scalability, all methods were evaluated at sample sizes of 50,000, 100,000, 150,000, 250,000, 350,000, and 450,000 individuals. Benchmarking was performed on chromosome 1, comprising 388,017 variants and 11,724 gene-tissue pairs from prediction models spanning 32 tissue and single-cell types. The prediction weight panel was obtained from our recent publication (Venkatesh et al., 2024), models used for benchmarking can be found in the Supplement Data. All methods were evaluated on identical genotype inputs. When comparator workflows required additional method-specific preprocessing or postprocessing steps, these steps were excluded from performance evaluation in favor of comparator methods. All jobs were run on the Mount Sinai Minerva high-performance computing environment under IBM Spectrum LSF (Load Sharing Facility) using CPU-only execution in the same containerized software environment (`marvany/ubuntu-r422-PLINK2:1.1`), with 1 CPU core allocated per job.

### Benchmark evaluation metrics

Output concordance between GROMTools and each reference method was assessed by sampling predicted expression values for different sets of 10,000 individuals per gene-tissue pair and performing pairwise comparisons across methods. For each comparison, we calculated Pearson correlation and root mean square error (RMSE) across all gene-tissue pairs, and reported the minimum Pearson correlation and maximum RMSE observed. Because the methods are deterministic, any residual discrepancies are attributable to minor floating-point rounding differences. Computational performance was evaluated using total computational hours and peak random-access memory (RAM) usage as reported by the LSF scheduler.

## Benchmark

### GROMTools preserves GReX estimates

Across all genes and tissues for our gromtools predictions, minimum Pearson’s correlation was greater than 0.999 and maximum RMSE was less than 0.001 when compared to both PrediXcan and PLINK2 GReX predictions.

### CPU-hours and peak memory usage decrease substantially across cohort sizes

Across the different sample subsets, when using GROMTools CPU time and peak memory usage decrease by ∼99% (107-fold against PrediXcan and 165-fold against PLINK2) and ∼96% (∼33-fold) respectively when compared to either PrediXcan or PLINK2 (**Fig. 1**) on average across sample sizes (**Supplementary Table 1** summarizes these performance metrics for each sample subset across methods).

**Fig. 1.**
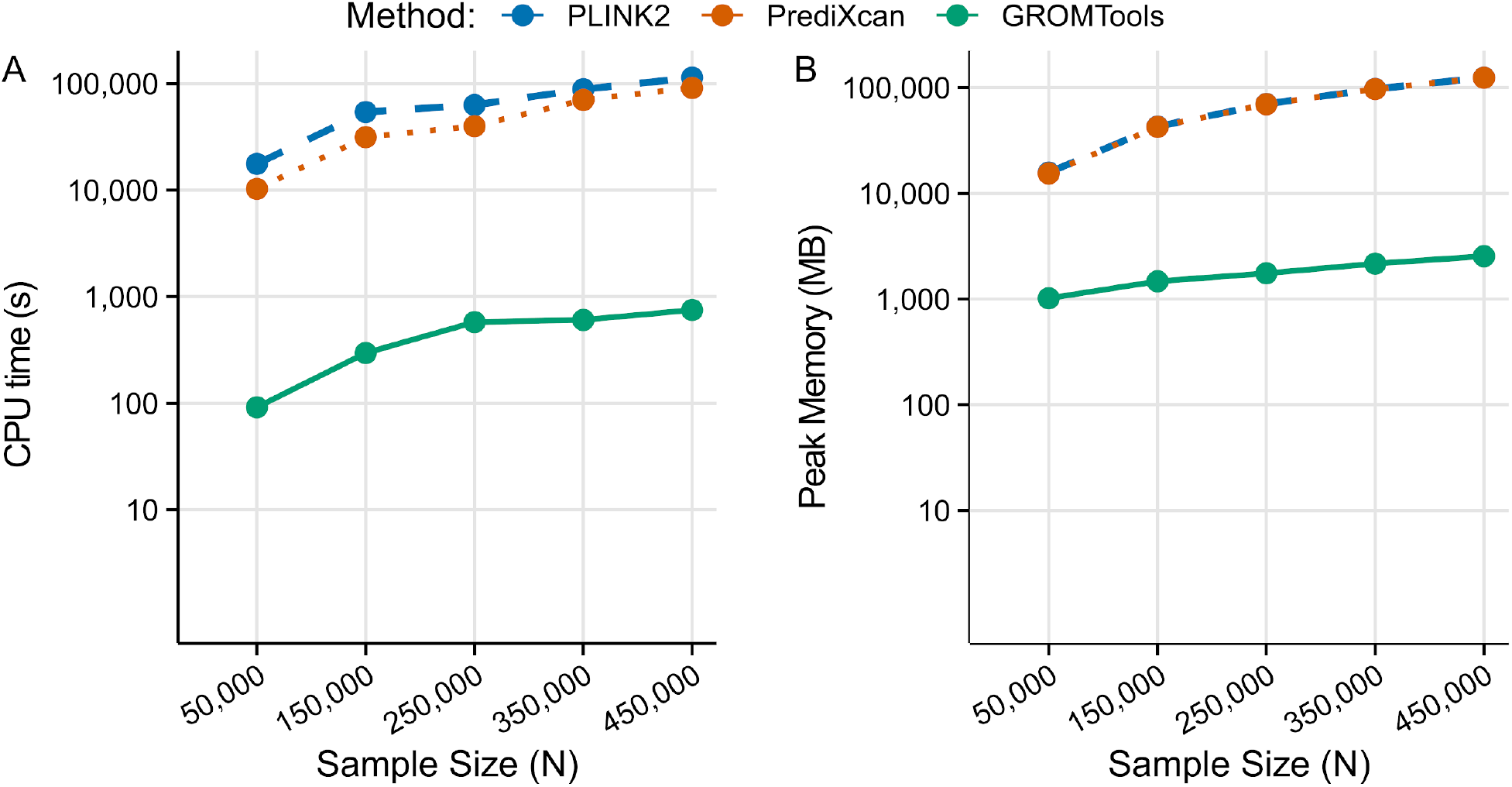
Scaling of CPU time and peak memory usage with sample size. **(A)** Total CPU time (seconds; log10 scaling) for GROMTools, PLINK2, and PrediXcan across sample sizes from 50,000 to 450,000 individuals for chromosome 1. GROMTools consistently required less CPU time than both comparator methods, with CPU-time reductions ranging from 108.93-fold to 208.75-fold relative to PLINK2 and from 69.02-fold to 121.39-fold relative to PrediXcan. **(B)** Peak memory usage (MB; log10 scaling) across the same sample sizes for chromosome 1. GROMTools maintained a smaller peak memory footprint, with peak memory reductions ranging from 15.46-fold to 48.82-fold relative to PLINK2 and from 15.13-fold to 48.63-fold relative to PrediXcan. These results highlight the substantially lower computational burden of GROMTools and its improved scalability for biobank-scale GREX imputation.

### Reduced memory and I/O burden enables practical biobank-scale GREX imputation

grom`_impute`’s binary output format reduces I/O overhead relative to the text-based outputs produced by comparator methods. For large numbers of genes or models, such workflows can generate hundreds of gigabytes of output, often requiring chunked execution and subsequent merging or format-conversion steps. Across benchmark sample sizes, this was reflected in lower I/O activity for GROMTools, with average read-character count (`rchar)` reduced by approximately 32-fold versus PLINK2 and 13-fold versus PrediXcan, and average write-character count (`wchar)` reduced by approximately 6-fold and 3-fold, respectively (**Supplementary Table 2**). The binary outputs can be read directly into downstream analyses in R, reducing storage demands and simplifying biobank-scale individual-level GReX workflows.

## Conclusions

GROMTools addresses a practical bottleneck in individual-level GREX imputation by reducing computational cost and memory requirements while preserving near-identical numerical results relative to baseline workflows. Across UK Biobank-scale benchmarks, the GROMTools reduces CPU-hours and peak memory by 99% (>100-fold) and 96% (∼33-fold), respectively, supporting routine large-scale GReX estimation on standard CPU-based compute environments. These gains were accompanied by lower I/O burden, as GROMTools streams directly from PLINK files and avoids large text-based score outputs, instead generating a compact binary format that can be streamed directly in R or loaded in chunks for downstream analyses.

Beyond computational efficiency, GROMTools simplifies workflow execution and, although it internally uses tabular files to improve readability and facilitate modification of transcriptomic imputation models, it also supports importing commonly used database formats such as the SQLite `.db` models established by PrediXcan. The method also produces a single unified output across all evaluated models, avoiding the chunking and post hoc merge steps often required in conventional high-dimensional imputation pipelines, where intermediate outputs can reach hundreds of gigabytes. Together, these features reduce pipeline complexity and improve usability in transcriptome-informed analyses.

Several limitations remain. First, the current implementation does not perform explicit allele alignment across model identifiers and therefore assumes consistent effect-allele orientation for overlapping variants with the provided QC scripts, across loaded models. Second, although GROMTools is efficient in total CPU-hours, the current version does not yet support parallel execution available in alternative implementations, and further reductions in wall-clock time will be achievable with future parallelization. Future development will focus on explicit allele-alignment support, expanded parallel execution with broader multithreading.

## Data Availability

All data used in this study are available from UK Biobank via its standard access procedures. UK Biobank data are available to eligible bona fide researchers for health-related research in the public interest through the UK Biobank Access Management System.

https://voloudakislab.github.io/gromtools/

https://doi.org/10.5281/zenodo.19547333

## Author contributions statement

M.A. and G.V. designed the experiment. M.A. coded the tool, M.A. and S.V. executed data analysis. All authors analysed, and discussed the results. M.A. and G.V. wrote the manuscript with input from all authors.

## Acknowledgements and funding

This study was also supported by the National Institutes of Health (NIH), Bethesda, MD under award numbers R01AG078657 (GV) and K08MH122911 (GV). This work was supported in part through the computational and data resources and staff expertise provided by Scientific Computing and Data at the Icahn School of Medicine at Mount Sinai and supported by the Clinical and Translational Science Award (CTSA) grant UL1TR004419 from the National Center for Advancing Translational Sciences.

## References

Gamazon, E.R. et al. (2015) A gene-based association method for mapping traits using reference transcriptome data. Nat. Genet., 47, 1091–1098.

Gusev, A. et al. (2016) Integrative approaches for large-scale transcriptome-wide association studies. Nat. Genet., 48, 245–252.

Huckins, L.M. et al. (2020) Analysis of Genetically Regulated Gene Expression Identifies a Prefrontal PTSD Gene, SNRNP35, Specific to Military Cohorts. Cell Rep., 31, 107716.

Krissaane, I. et al. (2020) Scalability and cost-effectiveness analysis of whole genome-wide association studies on Google Cloud Platform and Amazon Web Services. J. Am. Med. Inform. Assoc., 27, 1425–1430.

McDonald, S.A. et al. (2012) Fee-for-service as a business model of growing importance: the academic biobank experience. Biopreserv. Biobank., 10, 421–425.

Sudlow, C. et al. (2015) UK biobank: an open access resource for identifying the causes of a wide range of complex diseases of middle and old age. PLoS Med., 12, e1001779.

Venkatesh, S. et al. (2024) Single-nucleus transcriptome-wide association study of human brain disorders. medRxiv, 2024.11.04.24316495.

Voloudakis, G. et al. (2022) A translational genomics approach identifies IL10RB as the top candidate gene target for COVID-19 susceptibility. NPJ Genom Med, 7, 52.

Zeng, L. et al. (2024) A Single-Nucleus Transcriptome-Wide Association Study Implicates Novel Genes in Depression Pathogenesis. Biol. Psychiatry, 96, 34–43.

Zhang, W. et al. (2019) Integrative transcriptome imputation reveals tissue-specific and shared biological mechanisms mediating susceptibility to complex traits. Nat. Commun., 10, 3834.

