## Supplement Data, Supplementary Table 1, Supplementary Table 2, Supplementary Table 3 for "GROMTools: scalable individual-level GReX imputation for mega-biobank-scale cohorts"

Affiliations:
[1]Department of Psychiatry, Icahn School of Medicine at Mount Sinai, New York, NY, USA
[2]Center for Disease Neurogenomics, Icahn School of Medicine at Mount Sinai, New York, NY USA
[3]Friedman Brain Institute, Icahn School of Medicine at Mount Sinai, New York, NY, USA
[4]Department of Genetics and Genomic Science, Icahn School of Medicine at Mount Sinai, New York, NY, USA
[5]Mental Illness Research, Education, and Clinical Center (VISN 2 South), James J. Peters VA Medical Center, Bronx, NY, USA
[6]Center for Precision Medicine and Translational Therapeutics, James J. Peters VA Medical Center, Bronx, NY, USA.
[7]University of Crete School of Medicine, University Hospital of Heraklion, Heraklion, Crete, Greece
[8]Department of Artificial Intelligence and Human Health, Icahn School of Medicine at Mount Sinai, New York, NY, USA

### SUPPLEMENTARY TABLES

#### Supplementary Table 1

| Method | Sample size | CPU time (sec) | Peak memory (MB) |
|---|---|---|---|
| **GROMTools** | 50,000 | 91.53 | 1,017 |
| **GROMTools** | 100,000 | 168.90 | 1,209 |
| **GROMTools** | 150,000 | 295.58 | 1,472 |
| **GROMTools** | 250,000 | 577.11 | 1,753 |
| **GROMTools** | 350,000 | 604.49 | 2,163 |
| **GROMTools** | 450,000 | 748.37 | 2,543 |
| **PLINK2** | 50,000 | 17,648.17 | 15,720 |
| **PLINK2** | 100,000 | 35,257.94 | 29,267 |
| **PLINK2** | 150,000 | 54,028.79 | 42,829 |
| **PLINK2** | 250,000 | 62,862.50 | 69,936 |
| **PLINK2** | 350,000 | 88,437.69 | 97,046 |
| **PLINK2** | 450,000 | 113,614.30 | 124,144 |
| **PrediXcan** | 50,000 | 10,275.93 | 15,383 |
| **PrediXcan** | 100,000 | 20,186.21 | 28,912 |
| **PrediXcan** | 150,000 | 31,367.30 | 42,458 |
| **PrediXcan** | 250,000 | 39,831.50 | 69,542 |
| **PrediXcan** | 350,000 | 70,195.68 | 96,611 |
| **PrediXcan** | 450,000 | 90,841.42 | 123,656 |

**Supplementary Table 1.** Comparison of CPU time and peak memory usage across benchmark sample sizes for GROMTools, PLINK2, and PrediXcan.

#### Supplementary Table 2

| Method | Sample Size | Read-like I/O bytes (rchar) | Write-like I/O bytes (wchar) |
|---|---|---|---|
| **GROMTools** | 50,000 | 8.80E+08 | 4.70E+09 |
| **GROMTools** | 100,000 | 1.75E+09 | 9.39E+09 |
| **GROMTools** | 150,000 | 2.62E+09 | 1.41E+10 |
| **PLINK2** | 50,000 | 3.16E+10 | 2.99E+10 |
| **PLINK2** | 100,000 | 5.54E+10 | 5.63E+10 |
| **PLINK2** | 150,000 | 7.92E+10 | 8.28E+10 |
| **PrediXcan** | 50,000 | 1.27E+10 | 1.37E+10 |
| **PrediXcan** | 100,000 | 2.43E+10 | 2.75E+10 |
| **PrediXcan** | 150,000 | 3.59E+10 | 4.12E+10 |

**Supplementary Table 2.** Read-like and write-like I/O volume across benchmark sample sizes for GROMTools, PLINK2, and PrediXcan.

### Supplementary Table 3

| **Model** |
|---|
| Excitatory Neuron |
| Inhibitory Neuron |
| Immune Cell |
| Mural Cell |
| Endothelial Cell |
| Astrocyte |
| Oligodendrocyte |
| Oligodendrocyte Progenitor Cell |
| Layer 2-3 Intratelencephalic Excitatory Neuron |
| Layer 3-5 Intratelencephalic Excitatory Neuron 1 |
| Layer 3-5 Intratelencephalic Excitatory Neuron 2 |
| Layer 3-5 Intratelencephalic Excitatory Neuron 3 |
| Layer 5-6 Near-Projecting Excitatory Neuron |
| Layer 5 Extratelencephalic Excitatory Neuron |
| Layer 6 Corticothalamic Excitatory Neuron |
| Layer 6 Intratelencephalic Excitatory Neuron 1 |
| Layer 6 Intratelencephalic Excitatory Neuron 2 |
| Layer 6B Excitatory Neuron |
| ADARB2 Inhibitory Neuron |
| Ivy cell (LAMP5 LHX6 Inhibitory Neuron) |
| Neurogliaform cell (LAMP5 RELN Inhibitory Neuron) |
| Basket Cell (PVALB Inhibitory Neuron) |
| Chandelier Cell (PVALB CHC Inhibitory Neuron) |
| Martinotti and Non-Martinotti cell (SST Inhibitory Neuron) |
| VIP Inhibitory Neuron |
| Microglia |
| Perivascular Macrophage |
| Adaptive Immune Cell |
| Pericyte |
| Smooth Muscle Cell |
| Vascular Leptomeningeal Cell |
| Excitatory Neuron |
| Inhibitory Neuron |

**Supplementary Table 3.** Transcriptomic imputation models included in the benchmarking analyses. Only European ancestry models were utilized. Source: Venkatesh et al., 2026.
